# Relative Contributions of Family History and a Polygenic Risk Score on COPD and Related Outcomes: COPDGene and ECLIPSE studies

**DOI:** 10.1101/2020.07.23.20160739

**Authors:** Matthew Moll, Sharon M. Lutz, Auyon J Ghosh, Phuwanat Sakornsalkopat, International COPD Genetics Consortium, Craig Hersh, Terri H Beaty, Frank Dudbridge, Martin D Tobin, Murray A Mittleman, Edwin K. Silverman, Brian D. Hobbs, Michael H. Cho

**Author notes:** Correspondence: Michael H. Cho, MD, MPH., Channing Division of Network Medicine, 181 Longwood Ave 4^th^ floor, Boston, MA 02115. Author Contributions: Study Design: Matthew Moll, Sharon Lutz, Auyon J Ghosh, Murray Mittleman, Edwin K Silverman, Brian D Hobbs, Michael H Cho. Acquisition, analysis, or interpretation of the data: Matthew Moll, Brian D. Hobbs, Michael H. Cho, Sharon Lutz, Murray Mittleman, Edwin K. Silverman. Critical revision of the manuscript for important intellectual content: All authors. Statistical analysis: Matthew Moll, Brian D. Hobbs, Michael H. Cho, Sharon Lutz, Murray Mittleman, Edwin K. Silverman. Obtained funding: Edwin K. Silverman, Michael H. Cho. The **ECLIPSE** study (NCT00292552; GSK code SCO104960) was funded by GlaxoSmithKline. The content is solely the responsibility of the authors and does not necessarily represent the official views of the National Heart, Lung, and Blood Institute or the National Institutes of Health. **Competing Interests**: EKS received grant support from GlaxoSmithKline and Bayer. MHC has received grant support from GlaxoSmithKline and Bayer, consulting fees from Genentech and AstraZeneca, and speaking fees from Illumina. Dr. Hersh reports grant support from Boehringer-Ingelheim, Novartis, Bayer, and Vertex, outside of this study. MDT receives grant support from GlaxoSmithKline and Orion. The other authors declared no conflicts of interest. Additional Funding details and acknowledgements can be found in the supplementary materials.

## Abstract

**Introduction:** Family history is a risk factor for chronic obstructive pulmonary disease (COPD). We previously developed a COPD risk score from genome-wide genetic markers (polygenic risk score, or PRS). Whether the PRS and family history provide complementary or redundant information for predicting COPD and related outcomes is unknown.

**Methods:** We assessed the predictive capacity of family history and PRS on COPD and COPD-related outcomes in European ancestry subjects from the COPDGene and ECLIPSE studies. We also performed interaction and mediation analyses.

**Results:** In COPDGene, family history and PRS were significantly associated with COPD in a single model (P_FamHx_ = 1.6e-12; P_PRS_ = 5.0e-92). Similar trends were seen in ECLIPSE. Area-under-the-receiver-operator-characteristic-curves (AUCs) for family history, PRS, and the combined predictors for COPD were 0.752, 0.798, and 0.803, respectively. The AUC for a model containing both family history and the PRS was significantly higher than models with PRS (p = 0.00035) or family history (p = 6.1e-29) alone. Both family history and PRS were significantly associated with BODE, SGRQ, and multiple measures of quantitative emphysema and airway thickness. There was a weakly positive interaction between family history and the PRS under the additive, but not the multiplicative scale (RERI = 0.48, p=0.04). Mediation analysis found 16.5% of the effect of family history on risk for COPD was mediated through the PRS [95% CI: 9.4%-24.3%].

**Conclusion:** Family history and the PRS provide complementary information for predicting COPD and related outcomes. Future studies can address the impact of obtaining both measures in clinical practice.

## Introduction

Chronic obstructive pulmonary disease (COPD) is characterized by fixed airway obstruction, and is a leading cause of morbidity and mortality worldwide ^1^. This disease primarily develops in the context of cigarette smoking or biomass fuel exposure. However, only a minority of smokers develop this disease ^2,3^. Certain individuals may have increased genetic susceptibility to developing COPD, and studies have estimated the proportion of COPD liability variance explained by genetic factors (i.e. heritability) to be approximately 40% ^4–6.^

Before the advent of molecular genotyping, family history was the main method to assess a familial contribution to risk of complex diseases (i.e. where both genetic and environmental factors influence risk). Family history is a composite measure of common and rare variants as well as shared environmental risk factors ^7^. Family history is a known risk factor for COPD, with a population attributable risk of ∼17% ^8^. Despite the clinical use of family history, there are several limitations to using family history as a risk factor for a complex disease. First, a report of family history depends on the disease occurring and being diagnosed in relatives (typically first degree relatives), and does not capture those at high risk for the disease who may have not yet developed severe disease or received a diagnosis ^7^. Second, those affected by disease often do not have any known family history of the disease ^9^. Third, family history is subject to recall bias and often is unknown or incorrect. Finally, family history can reflect shared environmental exposures as well as genetics.

Genome-wide association studies (GWASs) have identified numerous single nucleotide polymorphisms (SNPs) associated with low lung function levels and COPD ^10–14^, though these variants typically exert a small effect on disease risk and account for a small proportion of the phenotypic variability. Pooling thousands to millions of genetic variants of small effect into a composite genetic risk score can improve prediction ^10,13,14^. We recently developed a polygenic risk score (PRS) highly predictive for COPD, multiple CT imaging phenotypes, and patterns of lung growth and decline that are thought to be important in disease pathogenesis. The PRS added information to traditional risk factors (age, sex, pack-years of smoking), and increased predictive power for COPD ^15^.

The relationship between genetic risk scores and family history in predicting COPD is unclear. Recently, studies have assessed relative contributions of genetic or polygenic risk scores and family history for other complex diseases. Genetic risk scores improved prediction of incident coronary heart disease, and were reported to be independent of family history ^16,17^. In schizophrenia, a PRS and family history were found to provide complementary information; however, the PRS was found to interact with family history, and 17.4% of the effect of family history on schizophrenia was mediated through the PRS ^18^. These data suggest that family history and PRS can provide complementary information in complex disease, and the way family history interacts with or is mediated by genetic risk may vary amongst diseases.

We hypothesized that PRS and family history would provide complementary information for predicting COPD and related phenotypes. We additionally asked whether the relative impact of family history and PRS differed among these related phenotypes; whether PRS and family history interact on the multiplicative or additive scale; and how much of the effect of family history on risk to COPD is mediated through the PRS.

## Methods

### Study populations

#### COPDGene

We included participants from the COPDGene study, which has been previously described ^19^. Briefly, this study included 10,192 non-Hispanic white (NHW) and African American participants aged 45 to 80 years with ≥ 10 pack-years of smoking. We included only NHW individuals in this analysis as the PRSs were derived from a European ancestry population and thus have higher predictive performance in European compared to non-European populations.

#### ECLIPSE

The ECLIPSE study was a 3-year longitudinal study to identify surrogate endpoints in 2746 participants (2164 COPD cases) associated with disease progression and exacerbations in COPD ^20^. Participants were white, 40-75 years of age at enrollment, and had ≥ 10 pack-years of smoking history.

In both cohorts, baseline demographic, spirometry, chest CT imaging, family history, exacerbation frequency over the last 12 months, and number of severe exacerbations (requiring emergency room visits or hospitalizations) were recorded. A schematic of the study design is shown in Figure 1.

**Figure 1:**
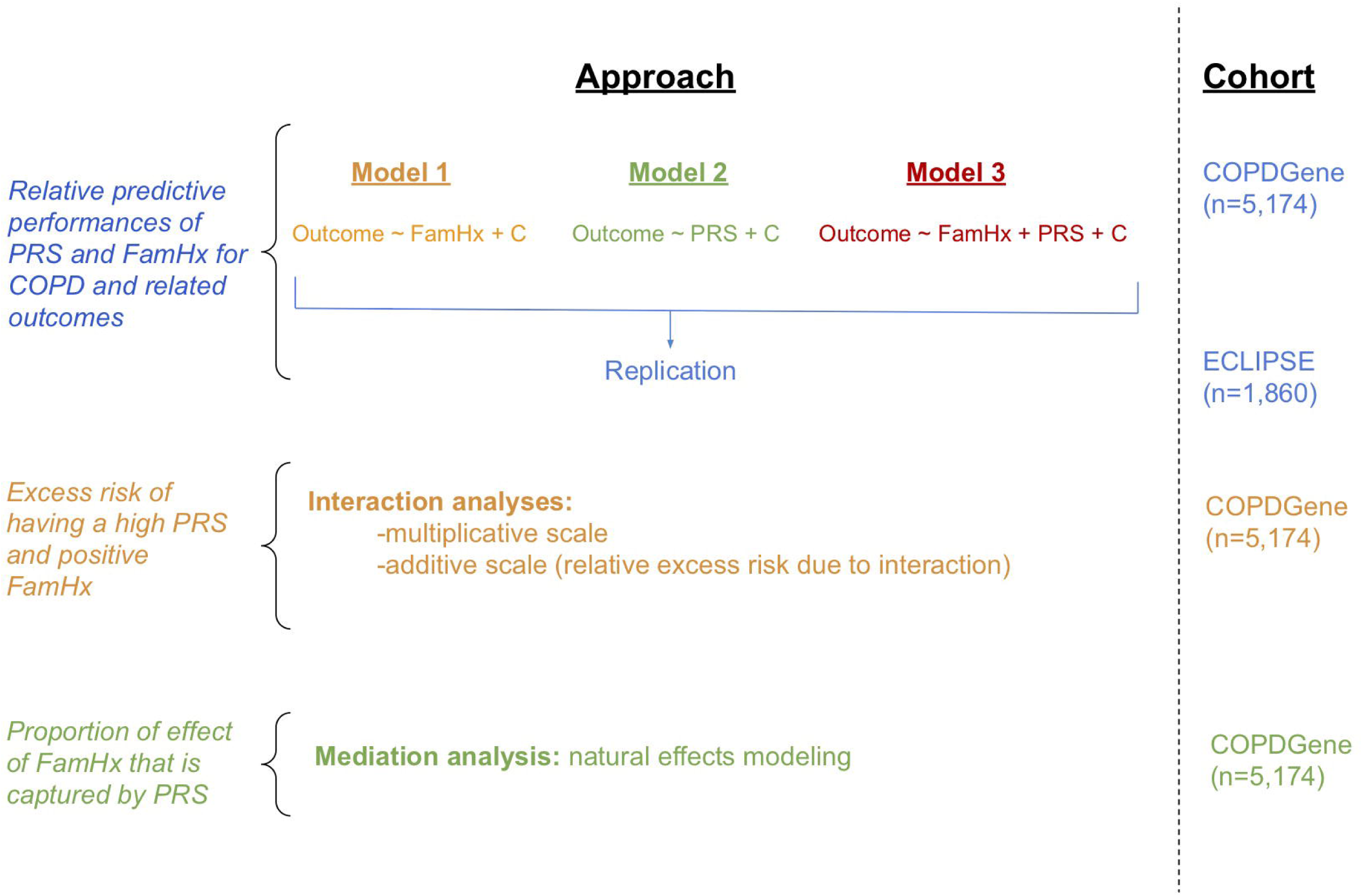
Schematic of study design.

### Statistical analyses

We utilized COPDGene as our discovery cohort and ECLIPSE as a replication cohort. The primary outcome was moderate-to-severe COPD, defined as post-bronchodilator forced expiratory volume in 1 second (FEV_1_) < 80% predicted and FEV_1_/forced vital capacity (FVC) < 0.7. Control subjects had FEV_1_ ≥ 80% predicted and FEV_1_/FVC ≥ 0.7; Global Initiative for Chronic Obstructive Lung Disease ^1^ (GOLD) 1 and preserved ratio with impaired spirometry (PRISm) subjects were excluded. Secondary outcomes included frequent exacerbations (>1 exacerbation in prior 12 months), severe exacerbations (exacerbation requiring hospitalization or an emergency room visit), death, baseline 6MWD (6-minute walk distance), the BODE index (body mass index (BMI), obstruction, dyspnea, exercise capacity) ^21^, total SGRQ (St. George’s Respiratory Questionnaire) score, and CT imaging phenotypes (percent emphysema determined by the percent low attenuation area of the lungs < −950 Hounsfield units [%LAA <-950 HU]^22^, the HU value at the 15^th^ percentile of the lung density histogram on inspiratory scans [Perc15] ^23^, square root of wall area of a hypothetical airway with an internal perimeter of 10 mm [Pi10] ^24^, mean wall area percent [WA %] ^22^). Additional outcomes only available in COPDGene included visual CT subtypes of destructive emphysema and airway pathology^25^. We adjusted for multiple comparisons using the Bonferroni method.

Consistent with prior analyses in the COPDGene study, we defined a family history of COPD by self-reported maternal or paternal history of COPD, chronic bronchitis, or emphysema^8^. All participants responded to the questionnaire; we considered a response of “Unknown” as a “No” response. In the ECLIPSE study there was not a question about family history of COPD, so a maternal or paternal family history of chronic bronchitis or emphysema was considered a positive family history for COPD. In ECLIPSE, there were no missing or “Unknown” responses.

Development of the PRS was previously described ^15^. Briefly, the PRS was derived from genome-wide association studies (GWASs) of FEV_1_ and FEV_1_/FVC ^13^. For each individual, we calculated a PRS for FEV_1_ a separate PRS for FEV_1_/FVC, then created a combined PRS using a weighted sum of these two scores. For the primary analysis, we treated the PRS as a continuous variable. We also dichotomized the PRS (1 = top tertile; 0 = bottom two tertiles) to allow comparison of odds ratios for the PRS with odds ratios for family history; tertiles were chosen as about one-third of controls also had a positive family history, so this dichotomization allowed comparison of the PRS to family history. The PRS was treated as a continuous variable for interaction and mediation analyses. We performed biserial correlation between family history and the PRS assuming 10 bins for the PRS using the polycor R package ^26^.

For both family history and the PRS (dichotomized), we calculated the odds ratio, attributable fraction in the exposed, and attributable fraction in the population with respect to the primary outcome as detailed in ^27^. We built three regression models (linear or logistic, as appropriate) for each outcome: Model 1) Outcome ∼ family history + covariates; Model 2) Outcome ∼ PRS + covariates; and Model 3) Outcome ∼ family history + PRS + covariates. All models were adjusted for age, sex, and pack-years of cigarette smoking. When frequent or severe exacerbations was the outcome, the model was additionally adjusted for baseline FEV_1_ % predicted and current smoking as these are known predictors of COPD exacerbations. For baseline 6MWD, models were adjusted for height and weight.

For death as the outcome, models were adjusted for the BODE index, which is a strong predictor of mortality in COPD ^21^. For all CT imaging outcomes, the models were adjusted for CT scanner. To estimate effects including all subjects, we meta-analyzed the results of models from COPDGene and ECLIPSE that included both PRS and family history (Model 3) using the meta R package ^28^. We constructed boxplots of odds ratios, betas, and 95% confidence intervals, as appropriate. We calculated the area-under-the-receiver-operator-characteristic-curve (AUC) for logistic regression models to assess predictive performance of each model. AUC and 95% confidence intervals were calculated with the pROC R package ^29^. For continuous outcomes, we calculated R^2^ and Akaike information criterion (AIC) values for each model. For each outcome, the model with the lowest AIC was considered the best fitting model.

For the primary outcome, moderate-to-severe COPD (hereafter, referred to as ‘COPD’), we performed an interaction analysis in the COPDGene study. We tested for a multiplicative interaction using a logistic regression model including family history, PRS, and a family history by PRS interaction term. We tested for an additive interaction by calculating the relative excess risk due to interaction (RERI)^27,30,31^. To test for mediated effects, we evaluated whether the COPD association with family history is mediated through the PRS by testing for the natural direct/indirect effect (NDE/NIE) ^32–34^ using Medflex ^35^ in R. Interaction and mediation models were adjusted for age, sex, and pack-years of cigarette smoking.

All analyses were done in R version 3.6.0 (www.r-project.org). Normality for continuous variables was assessed by visual inspection of histograms and Shapiro-Wilk tests. Results are shown as mean ± standard deviation or median [interquartile range], as appropriate. Differences in continuous variables were assessed with Student t-tests or Wilcoxon tests. Categorical variables were compared by ANOVA or Kruskal-Wallis tests, as appropriate.

## Results

### Characteristics of study populations

Characteristics of the study populations are shown in Table 1. We included 5,174 NHW participants from the COPDGene study (2,506 controls, and 2,668 COPD cases). COPD cases tended to be older, were more likely to be male, had greater mean pack-years of cigarette smoking, higher exacerbation rates, higher mean PRS, and a greater percentage of individuals reporting a family history of COPD.

**Table 1:** Characteristics of cohorts. Those with frequent exacerbations had more than one exacerbation per year requiring steroids and/or antibiotics. Severe exacerbations including worsening in respiratory health requiring emergency room visit or hospitalization.

|  | COPDGene NHW |  |  | ECLIPSE |  | *p*-value |
| --- | --- | --- | --- | --- | --- | --- |
|  | *Controls* | *Cases* | *p*-value | *Controls* | *Cases* |  |
| n | 2506 | 2668 |  | 147 | 1713 |  |
| Age in years (mean (SD)) | 59.54 (8.71) | 64.73 (8.13) | <0.001 | 57.32 (9.55) | 63.64 (7.10) | <0.001 |
| Sex (No. female, (%)) | 1283 (51.2) | 1187 (44.5) | <0.001 | 63 (42.9) | 563 (32.9) | 0.018 |
| BMI (kg/m<sup>2</sup>) (mean (SD)) | 28.83 (5.67) | 27.98 (6.02) | <0.001 | 27.34 (4.17) | 26.53 (5.54) | 0.115 |
| Current Smoking (No. (%)) | 1016 (40.5) | 912 (34.2) | <0.001 | 46 (37.7) | 475 (35.4) | 0.685 |
| Pack-years cigarette smoking (mean (SD)) | 38.78 (21.15) | 56.09 (27.27) | <0.001 | 31.01 (25.94) | 50.50 (27.47) | <0.001 |
| SGRQ Total Score (mean (sd)) (mean (SD)) | 17.05 (16.97) | 40.79 (20.86) | <0.001 | 9.45 (13.11) | 50.91 (19.89) | <0.001 |
| 6-minute walk distance (ft) (mean (SD)) | 1558.32 (330.46) | 1207.83 (388.93) | <0.001 | NaN (NA) | 1084.17 (355.24) | NA |
| BODE (mean (SD)) | 0.36 (0.80) | 2.96 (2.05) | <0.001 | NaN (NA) | 3.26 (2.13) | NA |
| Frequent Exacerbations (No<br>(%) > 1 per year) | 45 (2.5) | 214 (13.9) | <0.001 | 0 (0.0) | 775 (45.7) | <0.001 |
| Severe Exacerbations (No. (%)) | 63 (3.5) | 295 (19.2) | <0.001 | 0 (0.0) | 542 (31.6) | <0.001 |
| FEV1 % predicted (mean (SD)) | 94.85 (13.07) | 48.84 (17.80) | <0.001 | 108.87<br>(12.28) | 47.11 (15.47) | <0.001 |
| FEV1/FVC ratio (mean (SD)) | 0.77 (0.06) | 0.48 (0.13) | <0.001 | 0.80 (0.05) | 0.44 (0.11) | <0.001 |
| Combined FEV1 and FEV1/FVC<br>PRS (mean (sd)) (mean (SD)) | -0.27 (0.97) | 0.25 (0.96) | <0.001 | -0.60 (0.98) | 0.05 (0.98) | <0.001 |
| Family history of COPD, chronic<br>bronchitis, or emphysema (No.<br>(%)) | 710 (28.3) | 991 (37.1) | <0.001 | 51 (34.7) | 717 (41.9) | 0.108 |

In ECLIPSE, we included 1,860 European-ancestry participants, the majority of which were COPD cases (147 controls, 1,713 COPD cases). Differences between cases and controls were similar to those observed in the COPDGene sample, with the exception of current smoking status; COPDGene cases had a lower proportion of current smokers compared to controls, while ECLIPSE cases and controls had similar rates of current smoking. 6MWD, and therefore BODE, were not available in ECLIPSE controls.

### PRS and family history are complementary for predicting COPD

The distribution of PRS values in those with (n=1,701) and without (n=3,473) a family history of COPD in the COPDGene study is shown in Figure S1. Visual inspection reveals a small difference in the distribution of PRS values, though this difference was statistically significant (t-test p-value = 2e-7; biserial correlation coefficient = 0.093, p = 0.0062).

In each cohort, we considered three logistic regression models for moderate-to-severe COPD (see Methods, Figure 1). The resulting odds ratios for variables included in each model are shown in Table 2. When modeled separately in the COPDGene sample, holding all other covariates constant, family history was associated with a 1.77 odds ratio for COPD [95% CI: 1.6-2.0, p = 4.3e-17] and the PRS was associated with a 2.13 odds ratio for COPD [95% CI: 1.98-2.28, p= 9.0e-95] per standard deviation increment in the risk score. When modeled together, both family history and the PRS were associated with COPD (Family history: OR = 1.67 [95% CI: 1.45-1.92, p = 1.6e-12]; PRS: OR = 2.11 [95% CI: 1.97-2.27], p = 5.0e-92). In the ECLIPSE sample; when modeled alone (Model 1) family history was not significantly associated with risk of COPD, though when modeled along with PRS (Model 3), family history was significantly associated with risk of COPD (Table 2). Similar results were found when dichotomizing the PRS (Table S1)

**Table 2:**
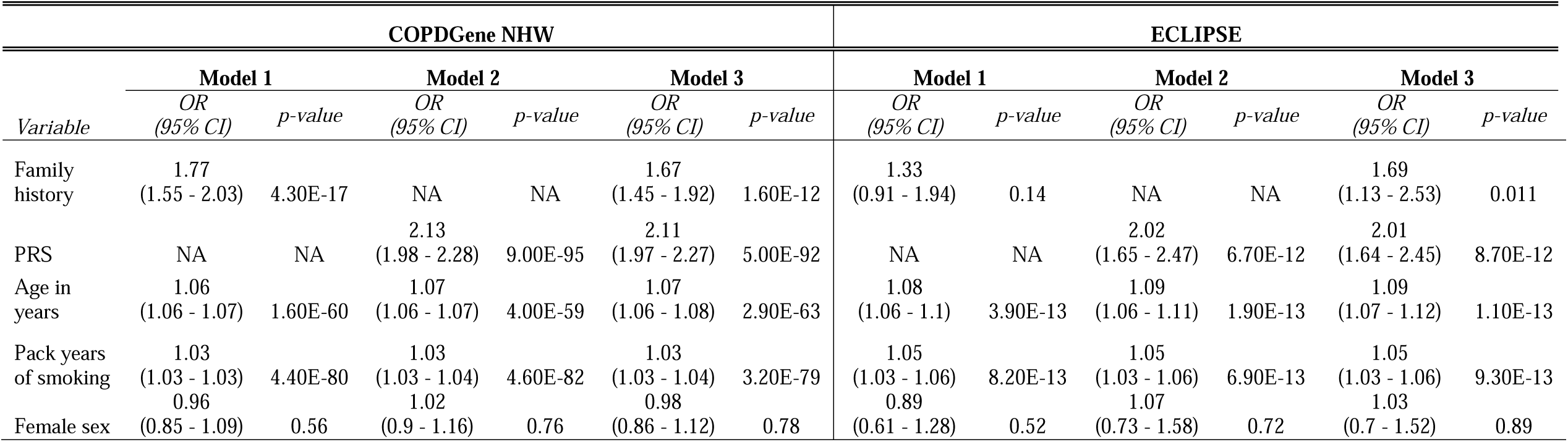
Associations of family history and PRS in three logistic regression models of moderate-to-severe COPD: Model 1 (COPD ∼ Family history + age + pack years + sex); Model 2 (COPD ∼ PRS + age + pack years + sex); Model 3 (COPD ∼ family history + PRS + age + pack years + sex). Bonferroni-adjusted level of significance is 0.05/3 models = 0.017. The PRS was treated as a continuous variable, and odds ratios are associated with 1 standard deviation increase in the PRS.

Utilizing COPDGene participants, we calculated the attributable fraction in the exposed (AF_exposed_) and in the population (AF_population_) for both family history and the dichotomized PRS (Table S2). The AF_exposed_ for family history was 0.37 and for the dichotomized PRS was 0.65. The AF_population_ for family history was 0.14, and for the PRS was 0.27.

### PRS and family history are complementary for predicting a range of COPD-related outcomes in COPDGene and ECLIPSE

We also considered three analogous models for a range of COPD-related outcomes (Table 3, Table S3). In COPDGene (Figure 2; Figure S2) AUCs for the prediction of COPD from family history (Model 1), PRS (Model 2), and the combined predictors (Model 3) were 0.752, 0.798, 0.803, respectively. The AUC for a model containing both family history and the PRS was significantly higher than models with only PRS (p = 0.00035) or only family history (p = 6.1e-29). For quantitative outcomes, we found models including both family history and PRS gave a better fit compared to those with PRS or family history alone, as evidenced by a lower AIC (Table S4). Family history was associated with frequent exacerbations (≥ 2 exacerbations per year) in COPDGene, and the PRS, but not family history, was significantly associated with a higher Pi10. Otherwise, both family history and the PRS were either concordantly associated even though some were not statistically significant.

**Table 3:** Association of family history and PRS with outcomes (*left*). All models had form Outcome ∼ family history + PRS + age + sex + pack years + C, where C equals any additional covariates listed in the table for a specific outcome. “CLE” = centrilobular emphysema. “BODE” = body-mass index, obstruction, dyspnea, exercise capacity index ^20^. SGRQ = St. George Respiratory Questionnaire. % LAA < −950 HU = percent low attenuation area of the lung less than −950 Hounsfield units. Perc15 = 15^th^ percentile of the lung density histogram on inspiratory scans. Pi10 = square root of wall area of a hypothetical airway with an internal perimeter of 10 mm. WA % = mean wall area percent. The PRS was treated as a continuous variable, and odds ratios are associated with 1 standard deviation increase in the risk score.

| Outcome | Covariates | COPDGene NHW |  |  |  | ECLIPSE |  |  |  |
| --- | --- | --- | --- | --- | --- | --- | --- | --- | --- |
|  |  | Family history<br>(OR (95% CI) or<br>beta (SE) |  | PRS (OR (95%<br>CI) or beta (SE) |  | Family history<br>(OR (95% CI) or<br>beta (SE) |  | PRS (OR (95% CI)<br>or beta (SE) |  |
|  |  |  | p |  | p |  | p |  | p |
| 6-minute walk distance | height, weight | -27 (11) | 0.016 | -33 (5.1) | 1.40E-10 | -29 (19) | 0.14 | 2.1 (9.2) | 0.82 |
| SGRQ Total Score |  | 0.27 (0.032) | 1.20E-16 | 0.14 (0.015) | 3.00E-21 | 3.6 (1.1) | 0.00084 | 2.2 (0.52) | 2.70E-05 |
| Frequent Exacerbations | FEV1 % predicted, current smoking | 1.65 (1.26 - 2.16) | 0.00031 | 0.91 (0.79 - 1.04) | 0.18 | 1.2 (0.94 - 1.52) | 0.15 | 0.99 (0.88 - 1.11) | 0.83 |
| Severe Exacerbations | FEV1 % predicted, current smoking | 1.17 (0.91 - 1.49) | 0.22 | 0.97 (0.86 - 1.1) | 0.66 | 0.96 (0.74 - 1.24) | 0.76 | 1.04 (0.93 - 1.18) | 0.48 |
| BODE |  | 0.39 (0.057) | 1.10E-11 | 0.35 (0.026) | 4.70E-39 | 0.13 (0.11) | 0.24 | 0.053 (0.053) | 0.32 |
| Dead | BODE | 1.15 (0.96 - 1.36) | 0.12 | 1.14 (1.05 - 1.24) | 0.0016 | 1.17 (0.94 - 1.44) | 0.16 | 1.01 (0.92 - 1.12) | 0.78 |
| % LAA < -950 HU | CT scanner | 0.37 (0.041) | 1.10E-19 | 0.19 (0.019) | 6.00E-24 | 0.42 (0.65) | 0.52 | 0.48 (0.3) | 0.11 |
| Perc15 | CT scanner | -6.1 (0.76) | 1.20E-15 | -3.4 (0.36) | 1.10E-21 | -2.4 (1.5) | 0.097 | -2.5 (0.68) | 0.00028 |
| Pi10 | CT scanner | 0.0092 (0.0039) | 0.017 | 0.013 (0.0018) | 2.30E-13 | -0.00037 (0.0089) | 0.97 | 0.0034 (0.0041) | 0.4 |
| WA % | CT scanner | 0.35 (0.093) | 0.00015 | 0.75 (0.043) | 5.00E-66 | 0.15 (0.22) | 0.5 | 0.82 (0.1) | 6.40E-16 |
| Paraseptal emphysema | CT scanner | 1.57 (1.17 - 2.11) | 0.0029 | 1.5 (1.29 - 1.74) | 6.20E-08 | NA | NA | NA | NA |
| Bronchial airway disease | CT scanner | 1.19 (0.83 - 1.71) | 0.34 | 1.64 (1.38 - 1.95) | 2.30E-08 | NA | NA | NA | NA |
| Small airway disease | CT scanner | 1.04 (0.68 - 1.57) | 0.87 | 1.79 (1.47 - 2.17) | 6.00E-09 | NA | NA | NA | NA |
| Mild CLE | CT scanner | 1.57 (1.22 - 2.03) | 0.00053 | 1.41 (1.25 - 1.6) | 4.20E-08 | NA | NA | NA | NA |
| Upper lobe CLE | CT scanner | 2.35 (1.46 - 3.77) | 0.00042 | 1.83 (1.42 - 2.35) | 2.50E-06 | NA | NA | NA | NA |
| Lower lobe CLE | CT scanner | 7.76 (2.19 - 27.41) | 0.0015 | 2.41 (1.3 - 4.47) | 0.005 | NA | NA | NA | NA |
| Diffuse CLE | CT scanner | 2.42 (1.64 - 3.57) | 9.10E-06 | 2 (1.63 - 2.45) | 3.70E-11 | NA | NA | NA | NA |
| Visual without<br>quantitative<br>emphysema | CT scanner | 2.12 (1.39 - 3.24) | 0.00051 | 1.65 (1.34 - 2.03) | 2.50E-06 | NA | NA | NA | NA |
| Quantitative without<br>visual emphysema | CT scanner | 1.66 (0.87 - 3.17) | 0.13 | 1.54 (1.11 - 2.13) | 0.01 | NA | NA | NA | NA |

**Figure 2.**
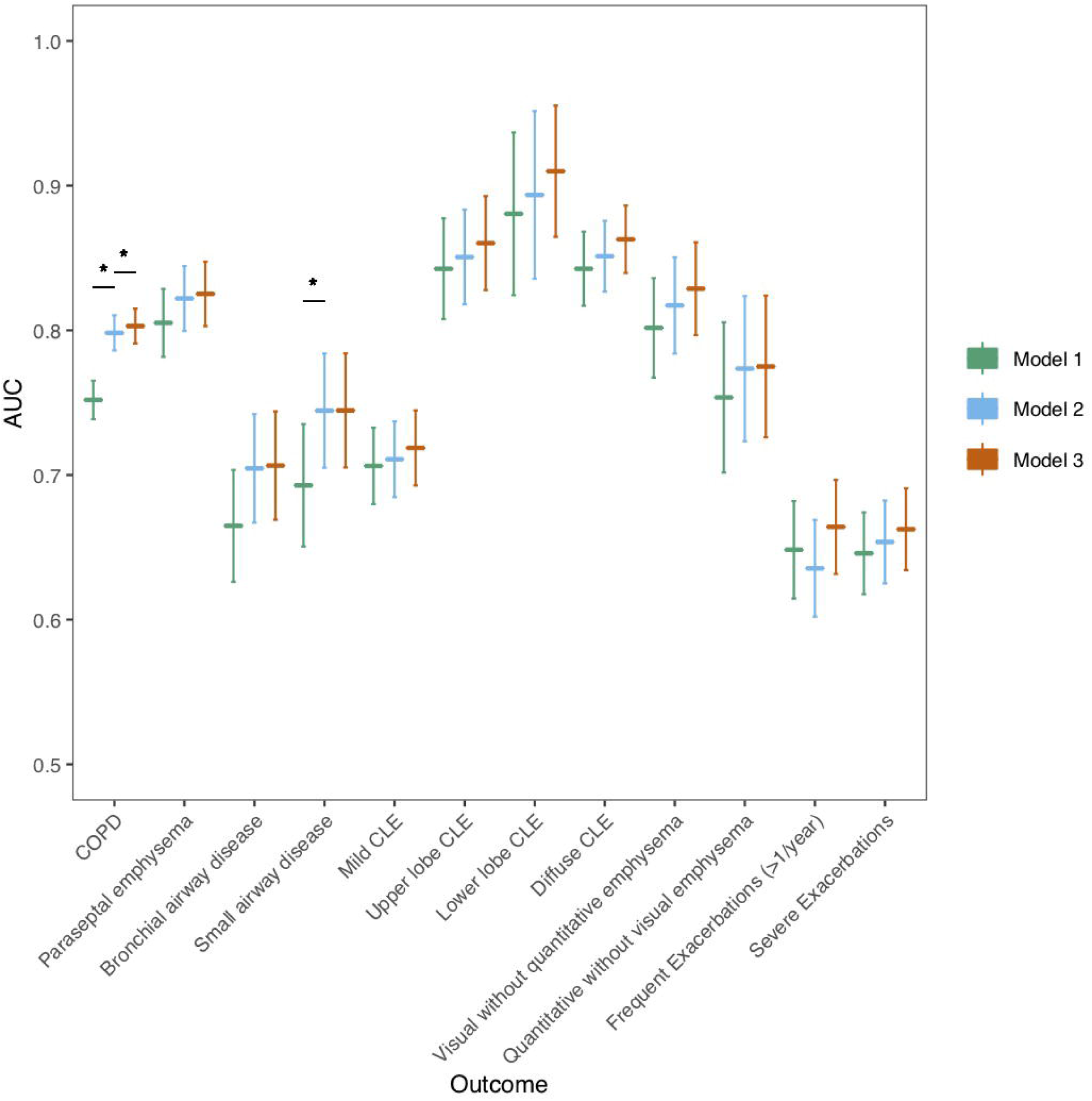
A) AUC analysis: Predictive performance (AUC) of three logistic regression models for outcomes show on the *x-axis* in the COPDGene study. The PRS was analyzed as a continuous variable. For each outcome, three models were trained: Model 1 (Outcome ∼ family history + age + sex + pack-years), Model 2 (Outcome ∼ PRS + age + sex + pack-years), and Model 3 (Outcome ∼ family history + PRS + age + sex + pack-years). “*” indicates that the p-value comparing model AUCs was less than Bonferroni-corrected level of significance (p < 0.05/12 = 0.0036). B) Predictive performance (R^2^) of three linear regression models for outcomes show on the *x-axis* in the COPDGene study. For each outcome, three models were trained: Model 1 (Outcome ∼ family history + age + sex + pack-years), Model 2 (Outcome ∼ PRS + age + sex + pack-years), and Model 3 (Outcome ∼ family history + PRS + age + sex + pack-years). Abbreviations are listed in the caption for Table 3.

We found similar results in ECLIPSE (Figure S3 and S4). Model 2 demonstrated significantly improved performance over Model 1, but Model 3 performed similarly to Model 2 for COPD prediction. For COPD-related traits, results were similar to COPDGene for all outcomes except for WA % and Pi10 in ECLIPSE. Results are also presented as a meta-analysis (Figure 3, Figures S5, S6, S7).

**Figure 3:**
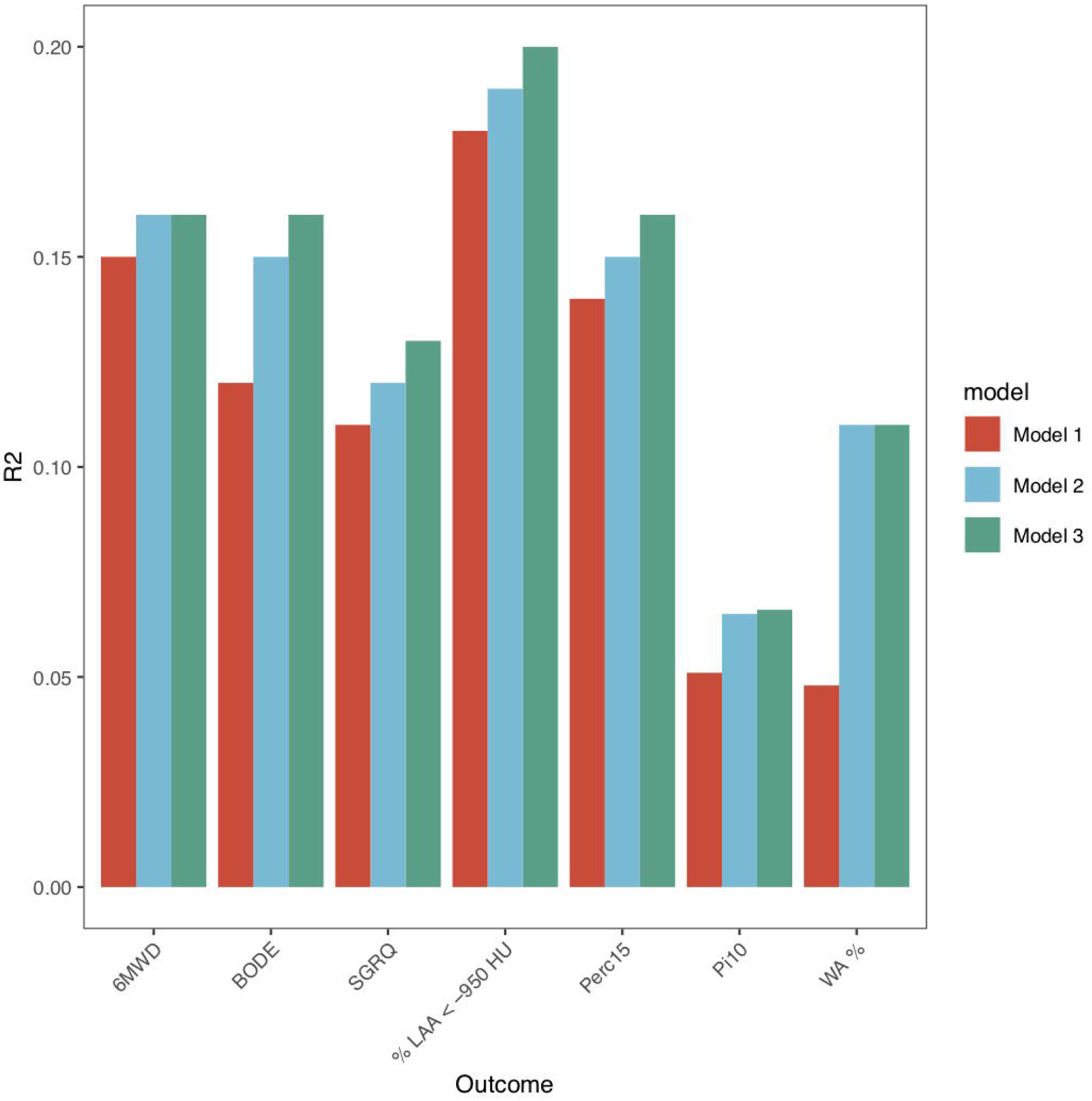
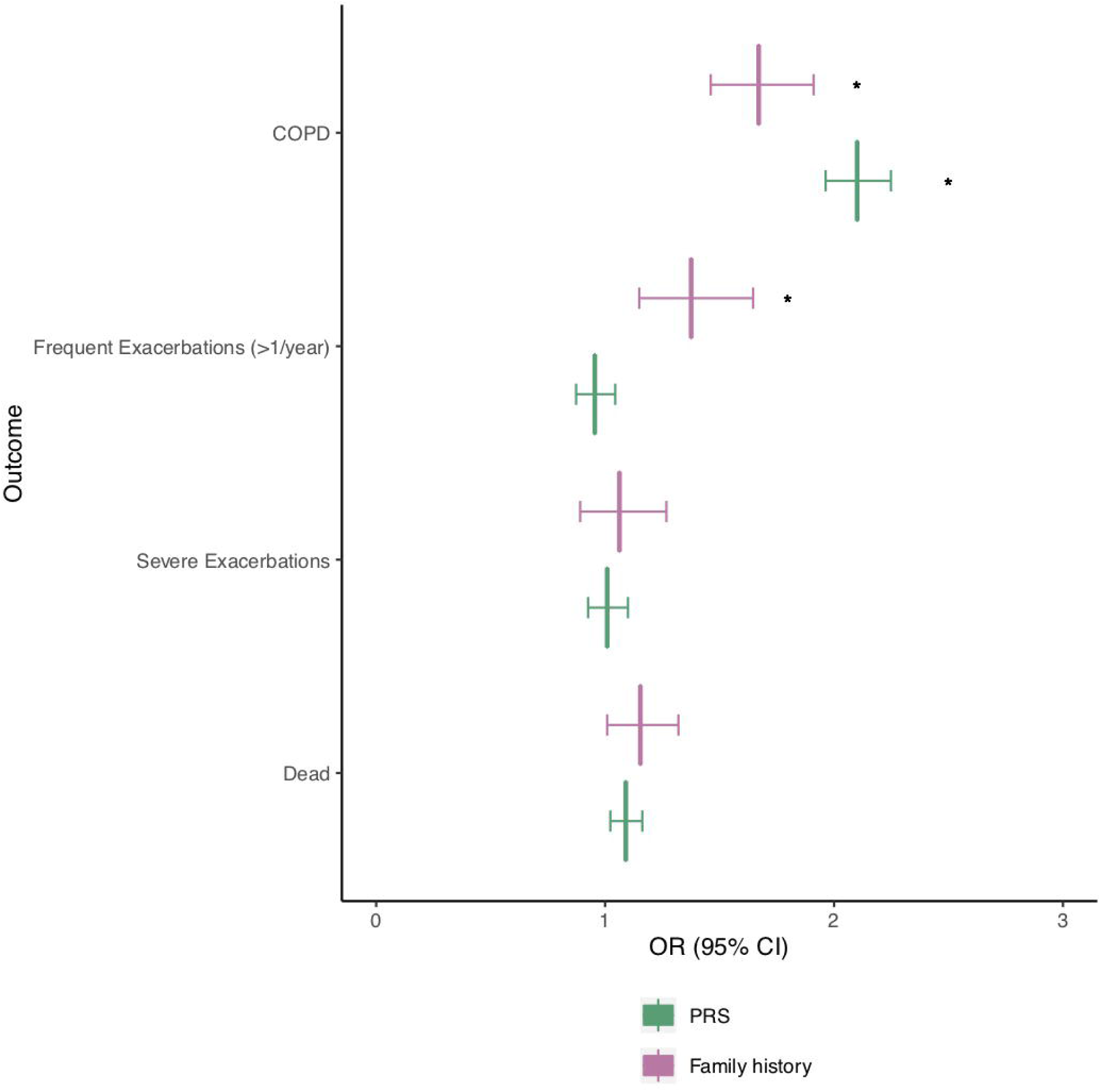
Meta-analyses of binary outcomes with PRS treated as a continuous variable. COPDGene and ECLIPSE studies were meta-analyzed, and fixed effects odds ratios with 95% confidence are shown for family history and PRS for each outcome. Odds ratios for the PRS indicate the odds ratio for the listed outcome for every standard deviation increase in the PRS. “*” indicates that p-value was less than Bonferroni-corrected level of significance (0.05/11 = 0.0045 (includes 7 continuous outcomes shown in supplement). Abbreviations are listed in the caption of Table 3.

### Analysis of interaction between family history and the PRS

As gene-gene and gene-environment interactions could lead to excess disease risk, we sought to understand whether having both a family history and higher PRS was associated with a greater odds ratio for COPD than would be explained by either risk factor alone. Using the COPDGene study, we investigated whether there is an interaction between family history and the PRS. The odds ratio for COPD for any given PRS value, separated by family history of COPD, is shown in Figure S8. In a logistic regression model of COPD, the family history * PRS interaction term was not significant (β = −0.09 (SE: 0.08), p=0.3) on the multiplicative scale. The RERI was 0.48 [95% CI: −0.04-1.00, p=0.04], suggesting some significant interaction on the additive scale.

### The effect of family history is partially mediated through PRS

We employed a natural effects model to investigate whether, and to what extent, the effects of family history are mediated through the PRS. The directed acyclic graph on which this analysis was based is shown in Figure S9. There was a significant natural direct effect of family history on COPD (β = 0.48, p = 2.0e-13) and a significant indirect effect of family history on COPD through the PRS (β = 0.09, p = 8.7e-6). The proportion of the effect of family history mediated through the PRS was 16.5% [95% CI: 9.4%-24.3%].

## Discussion

In this study of 7,034 participants from two COPD case-control cohorts, we compared the relative effects of family history and a polygenic risk score (PRS) on COPD and related clinical and chest CT quantitative imaging phenotypes. We observed that family history and PRS provide complementary information in association with COPD and COPD-related phenotypes. Moreover, we demonstrated that a PRS based on genotyping of SNPs is more predictive of COPD than family history, which is an important milestone for genetic predictors. We also observed a small interaction between these two predictors on the additive scale. We also showed approximately 17% of the effect of family history on COPD is mediated through the PRS. These data highlight the relative strengths and limitations of both family history and the PRS as predictors, and provide evidence that both genome-wide SNP genotyping and family history can contribute to an understanding of an individual’s risk for COPD. While comparative analyses of family history and PRSs have been performed in cardiovascular ^16,17^ and psychiatric diseases ^18^, to our knowledge, this is the first analysis of the relative contributions of family history and a PRS to COPD risk.

Our findings are consistent with prior reports of the effect of positive family history on risk to COPD ^8,36,37^. In a prior study in COPDGene (limited to the first 2,500 participants), a family history of COPD was associated with a 1.73 odds ratio for the disease ^8^. Our study included more individuals (5,174 cases and controls versus 1,597) and observed a similar effect size (OR=1.77 versus OR=1.73) with a similar population attributable risk (14% versus 17%). Consistent with reports that adoptees with at least one parent with COPD are more likely to have COPD than adoptees with no parents with COPD ^36^, we found that a significant proportion of the effect of family history is mediated through the PRS.

Our findings have several implications. First, our findings suggest that family history and the PRS provide complementary information about an individual’s risk for COPD. We observe a small, but statistically significant overlap in effects of family history and the PRS. This is supported by the fact that family history and the PRS had similar effect sizes before and after being combined into a single regression model. As family history must be a composite measure for genetics and shared environment, the residual effect of family history on COPD after adjusting for the PRS is presumably driven by shared environmental effects, or genetic factors not included or inaccurately modelled by the PRS. On the other hand, family history can only capture contributions from those whose parents developed and were aware of their diagnosis of disease, while the PRS is able to capture disease risk in the absence of any knowledge of parental disease. Our findings are also consistent with the current thinking that most complex disease cases are sporadic, occurring in individuals without a family history of the disease^9^, and emphasize the potential predictive value added by obtaining a PRS for individual patients.

Second, we observe that for COPD itself, and for certain related quantitative phenotypes, the PRS has more explanatory power than family history, and vice versa. The relative contribution of PRS versus family history depends on factors such as the relative contribution of the environment, the prevalence of the disease ^38^ and the accuracy of reported family history. The PRS had a larger effect size on risk to COPD affection status compared to family history. We estimated the attributable percent in the exposed for family history to be 37%, implying that if no one had a family history of COPD, 37% fewer cases would have occurred. By contrast, the PRS had a population attributable risk of 27% and attributable percent in the exposed of 65%. The PRS also had a larger association with BODE, and two measures of airway wall thickness (Pi10 and WA %), compared to family history. For COPD exacerbations, we previously demonstrated an association between the PRS and exacerbations, though this association did not persist after adjusting for baseline lung function level. Here, we confirm^8^ an association between family history and frequent exacerbations (i.e. ≥ 2 exacerbations in the previous 12 months), after adjusting for baseline FEV_1_ % predicted in the full COPDGene study, with a trend towards association between family history and frequent exacerbations in ECLIPSE. Whether this association represents shared environmental or genetic effects is not clear, particularly since PRS itself was not associated with frequent exacerbations. Only a few studies have examined genetic risk to COPD exacerbations, and no studies to our knowledge have studied the heritability of exacerbations. Future investigations into the genetics of COPD-related exacerbations and the role of shared environment may clarify these issues.

Third, we demonstrate family history is associated with risk to COPD through a direct effect as well as an indirect effect mediated via PRS. In addition, we report ∼17% of the effect of family history on COPD susceptibility is mediated through genetic variation, summarized by the PRS. Our finding is similar to the partial mediation of the effects of family history and a genetic risk score in predicting risk to schizophrenia ^18^. However, it is important to note that the indirect effect can be under-estimated if there is measurement error associated with the mediator (i.e. the PRS), and over-estimated if there is measurement error associated with the exposure (i.e. family history). Given the challenges of accurate diagnosis of COPD and the possibility of recall bias, we suspect family history may be particularly error-prone. Thus, the proportion of COPD risk due family history mediated through the PRS (16.5%) is likely on the lower end of our reported confidence intervals (95% CI: 9.4%-24.3%). Regardless, our analyses suggest that a large component of the effect of family history on COPD could be due to shared environmental factors.

We also observed an interaction between family history and the PRS on the additive scale (but not multiplicative scale). Thus, on an additive scale, those with both a family history and a high PRS are at higher risk than would be predicted by each risk factor independently. Though this interaction was statistically significant, the effect was modest and thus the interaction between family history and the PRS may not be clinically relevant.

Our study is the first to our knowledge that places family history of COPD in the context of a PRS. Compared to prior studies of family history in COPD, our sample size is larger and tests additional related phenotypes, including CT imaging. However, our study has several limitations. The study was performed in enriched cohorts of heavy smokers at high risk for COPD, and it is not clear what the benefit of collecting both family history and genotyping would be in the general population. Detailed questions about COPD history (which include COPD, chronic bronchitis and emphysema) are not routinely obtained in most population-based studies.

Family history is particularly susceptible to recall bias, and is often unknown or incomplete. Despite this, we observed significant effects of family history on multiple outcomes. Some of our findings failed to replicate in ECLIPSE potentially due to reduced power. All participants were smokers, making it difficult to tease out the contribution of shared environment on the effects of family history; however, future studies including never smokers could help address these issues. The PRS was based on common variants, and therefore, the contribution of rare variants to COPD risk and the overlap with family history was not assessed in the current study. Finally, there is arguably no proven strategy for prevention of COPD aside from smoking cessation, even in high-risk individuals (based on genetic markers). Potentially, certain high-risk occupations could be avoided. Further investigations are required to address the clinical impact of knowing one’s family history and measured genetic risk for COPD.

In conclusion, we demonstrate how family history and the PRS provide complementary information for predicting COPD and related phenotypes, though the PRS appears to be a stronger predictor than family history alone. Future studies should examine the impact of using information from both measures on clinical practice.

## Data Availability

No new data were generated in this study. Weights and R code for polygenic risk scores were previously reported and are publicly available at http://www.copdconsortium.org/announcements/10polygenicriskscorercode.

